# Chronic adaptive versus conventional deep brain stimulation in Parkinson’s disease: a blinded randomized pilot trial

**DOI:** 10.1101/2025.02.20.25322374

**Authors:** Ioannis U. Isaias, Sara Marceglia, Linda Borellini, Enrico Mailland, Filippo Cogiamanian, Sergio Barbieri, Antonella Ampollini, Elena Pirola, Luigi Remore, Laura Caffi, Chiara Palmisano, Claudio Baiata, Salvatore Bonvegna, Luigi Romito, Roberto Eleopra, Vincenzo Levi, Anna Rita Bentivoglio, Carla Piano, Alessandro Izzo, Maurizio Zibetti, Leonardo Lopiano, Michele Lanotte, Tomasz Mandat, Filippo Tamma, Mattia Arlotti, Costanza Conti, Lorenzo Rossi, Guglielmo Foffani, Andres Lozano, Elena Moro, Jens Volkmann, Marco Locatelli, Alberto Priori

## Abstract

**Background:** Adaptive deep brain stimulation (aDBS) is becoming a feasible therapeutic option in patients who are candidates for DBS, and implantable devices are now commercially available. We present the results of a blinded, randomized, crossover pilot trial aimed at comparing aDBS with conventional DBS (cDBS).

**Methods:** Fifteen patients were implanted with the AlphaDBS device (Newronika SpA, Milan, Italy, NCT04681534). The device replaced a previous Medtronic Activa PC at battery depletion in 11 patients, while the other four were first-time implant patients. Patients underwent two study phases: 1) a two-day, short-term follow-up in the hospital, in which patients received aDBS and cDBS for one day each (in random order); 2) a one-month, long-term follow-up at home, with patients receiving both DBS modes, each for two weeks. Safety endpoints were the occurrence of stimulation-related adverse events and the total electrical energy delivered to the tissue. The clinical endpoints were the Unified Parkinson’s (part III) and Dyskinesia Disease Rating Scales used for in-clinic evaluation, and a three-day diary used for home assessment to estimate good on time without troublesome dyskinesia. At the end of the study, patients blindly decided their preferred stimulation mode.

**Findings:** No related adverse events were reported. The AlphaDBS device reliably recorded deep brain signals and applied a linear algorithm that changed the stimulation current every minute based on the average local field potential amplitude, calculated in a patient-specific beta frequency range. In the short-term follow-up, aDBS improved patients as much as cDBS, with lower UDysRS scores. In the long-term follow up, considering intra-patient differences, aDBS provided greater benefit than cDBS in 80% of patients. The same percentage of patients preferred and continued with aDBS (mean follow-up 316 days).

**Interpretation:** Our results suggest that aDBS is safe and effective and can be applied in a large population of parkinsonian patients who are candidates for DBS. The majority of patients improved more with aDBS than cDBS, who subjectively preferred aDBS in the long term. Further research is needed to better understand the profile of the best responders and the scheduling of aDBS.

## INTRODUCTION

Almost two decades after its first conceptualization ^1^, adaptive deep brain stimulation (aDBS) has finally reached the level of clinical investigation for Parkinson’s disease (PD). In contrast to conventional DBS (cDBS), in which stimulation is constantly active with fixed parameters, stimulation in the closed-loop adaptive approach is changed in real time to respond to the patient’s symptoms and promptly deliver the optimal amount of therapy ^2–4^.

The adaptive approach is based on two major technological principles: the “biomarker” (i.e., the control variable that is used to estimate the patient’s symptoms) and the algorithm that translates changes in the biomarker into a new set of stimulation parameters ^5,6–8^. The biomarker used in devices capable of delivering aDBS is the activity (i.e., local field potentials, LFPs) recorded by the electrode chronically implanted in the subthalamic nucleus (STN), with specific focus on oscillations in the beta range (13-35 Hz) that parallel akinetic-rigid symptoms and levodopa-dependent brain states ^5,9^.

Currently available algorithms are based on patient-specific beta power thresholds. The “single threshold” algorithm (Percept PC, Medtronic Inc.) enables rapid ramping between a minimum and maximum current when the beta power exceeds a certain level for a given time ^10,11^. The “dual threshold” algorithm (Percept PC, Medtronic Inc.) features slow modulation of the delivered current, aiming to confine beta oscillations between two beta power levels. The “linear proportion” algorithm (AlphaDBS, Newronika SpA) used in our study instead provides timely modulation of the stimulation current within two beta power levels ^6,12–14^.

Whatever the strategy, aDBS brings high expectations in terms of increased benefits to patients, better understanding of patient responses to stimulation, and improved control of long-term changes in disease manifestation and stimulation-induced adverse effects ^15,16^. In addition, the possibility of continuous recordings of deep neuronal signals opened the way to the collection of neurophysiological “big data” that will allow knowledge on DBS mechanisms and its interaction with brain functions to be deepened, fostering precise electrical brain therapy.

For these reasons, conducting clinical investigations into aDBS is now crucial not only to demonstrate its safety and efficacy, but also to define new programming guidelines and start collecting chronic data to develop new algorithms for the analysis and interpretation of the data.

Accordingly, we present the results of a double-blind, randomized clinical investigation that aimed to assess the safety and efficacy of aDBS versus cDBS in patients with PD ^17^.

## METHODS

### Study design

The full protocol has been published previously ^17^ and the study is registered at clinicaltrials.gov (trial registration number: NCT04681534). In summary, this is a multicenter, randomized, crossover study in which cDBS was used as control. The study protocol was organized in two phases: the “short-term follow-up” (ST-FUP) and the “long-term follow-up” (LT-FUP). In the ST-FUP, randomized patients underwent two days of experimental sessions (i.e., one per each type of stimulation mode, cDBS and aDBS), in a controlled environment (i.e., during hospitalization). In the LT-FUP, patients deemed suitable by the neurologist continued at home with the AlphaDBS device, delivering aDBS or cDBS for two weeks in each mode, according to the randomization schedule.

This is a pre-market study that was first conducted under the EU-MDD regulation and then switched to the EU-MDR regulation in its last period. The full description of the AlphaDBS system, of the types of stimulation, and of the LFP sensing technology has been reported previously ^6,17^.

### Participants

The study initially included patients with PD chronically treated with cDBS and in need of a replacement of the implantable pulse generator (IPG; IPG replacement group), and was extended to patients with PD at implant (de novo group). Six investigational sites were included (four in Italy, one in The Netherlands, and one in Poland). We first recruited parkinsonian patients already on cDBS treatment because we already knew the chronic benefit from optimized bilateral STN-DBS. These patients were accustomed to cDBS and would have been able to identify treatment-associated adverse events or changes in efficacy. Additionally, the recorded STN-LFP oscillations in these patients would not be affected by the micro-lesioning effect that occurs after electrode implantation (stun effect).

Patients at IPG replacement were included in the study if treated with bilateral STN-DBS using a Medtronic Activa PC or Activa RC IPG (mono-channel or dual channel), had an active STN-DBS for at least three years, and were in need of IPG replacement within one year after consent. The de novo group consisted of patients who needed STN-DBS independently of this study and were selected according to the Core Assessment Program for Surgical Interventional Therapies (CAPSIT) guidelines (CAPSIT-PD) ^18^. These patients received quadripolar leads (Model 3389, Medtronic Inc, Minneapolis, USA) and the AlphaDBS IPG on the same day. All inclusion and exclusion criteria and study site information are available in the published study protocol ^17^.

The study was carried out in accordance with the Declaration of Helsinki, as amended by the 64th General Assembly of the World Medical Association, Fortaleza, Brazil, October 2013. The study obtained Ethical and Administrative approvals from all investigational sites and from Competent Authorities in Italy, Poland, and The Netherlands.

### Randomization and masking

Randomization was managed by an external entity (Contract Research Organization), according to the procedure described in the study protocol ^17^. Both the patient and clinicians in charge of patient assessment were unaware of the type of stimulation for the entire duration of the follow-up. The type of stimulation used in the first two weeks of the LT-FUP (aDBS or cDBS) was determined by the randomization schedule of the ST-FUP. At the end of the trial, patients were still blinded when asked for their preference between the two stimulation modes.

### Procedures and assessments

The detailed description of the study procedures can be found in the study protocol ^17^ and are summarized in Figure 1. In short, after screening, eligible patients underwent surgery (either for IPG replacement or implant). IPG replacement patients directly entered the ST-FUP, whereas de novo patients waited approximately four to five weeks (healing phase) with stimulation off but active sensing. The ST-FUP consisted of two experimental days (Day 1 and Day 2) preceded by up to five days of programming in cDBS or aDBS for fine-tuning of the stimulation, medications, and beta threshold adjustment. Each experimental day started with a baseline evaluation in meds-off/stim-off (T0), after overnight withdrawal of dopaminergic medications and at least thirty minutes after pausing STN-DBS. The subsequent follow-up evaluations were in a) meds-off/stim-on, after activating DBS for about one hour in either conventional or adaptive mode (T1), b) meds-on/stim-on, after one hour (T2) and six hours (T3) from the intake of levodopa and active STN-DBS, c) in meds-off/stim-on (T4), the day after, before restarting the experimental procedure. At all time points, we collected the Unified Parkinson’s Disease Rating Scale (UPDRS) motor part (part III), and the Unified Dyskinesia Rating Scale (UDysRS). The LT-FUP consisted of one month in which the patient received each stimulation mode for two weeks, according to the randomization schedule, and started immediately after the ST-FUP. To measure good-on-time (GOT) during the LT-FUP, patients were instructed to complete a three-day diary ^19^, in which they recorded their status (sleep, OFF, ON without dyskinesias, ON with mild, moderate, or severe dyskinesia) every thirty minutes. At the end of the first two weeks, patients were assessed in clinic with the UPDRS-III and UDysRS in the best-on state, after the intake of the morning medications and with active STN-DBS, before changing the stimulation mode. At the end of the LT-FUP, the patient decided whether to keep the AlphaDBS system in the preferred mode (cDBS or aDBS) (open-label extension phase) or to switch to a commercial IPG. During the open-label extension phase, patients received routine visits to assess system safety and efficacy. The pharmacological therapy was kept constant during the ST-FUP and LT-FUP, but was adjusted according to clinical routine in the open-label extension phase.

**Figure 1.**
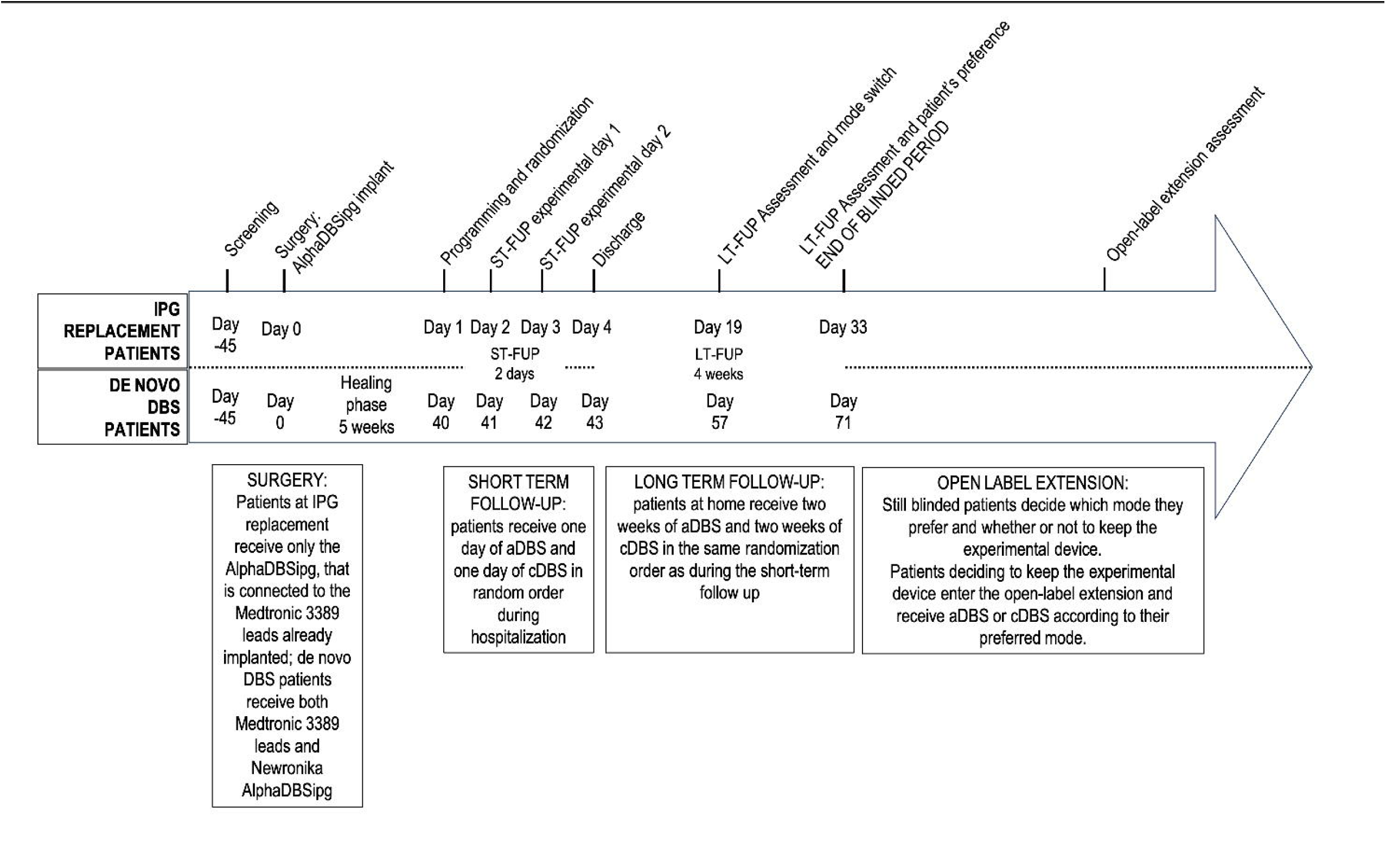
Study design. The diagram represents the sequence of study visits (top row) and phases (arrow) for patients at IPG replacement and de novo DBS patients. Abbreviations: (a/c)DBS, (adaptive/conventional)deep brain stimulation; IPG, implantable pulse generator; LT/ST-FUP, long-term/short-term follow-up.

### Adaptive programming

The AlphaDBS is a rechargeable device capable of recording LFP directly from the chronically implanted electrodes. Continuous recordings are stored in a memory located inside the IPG. Data are downloaded in the patient recharger at each recharging session and stored in the WebBioBank system ^6^.

The average LFP amplitude calculated in a patient-specific beta frequency range and normalized for the total amplitude in the 5-34 Hz range is used by the AlphaDBS system to drive the linear proportional algorithm that changes the stimulation current every minute. The averaging procedure consists of an exponential moving average with a time constant of fifty seconds over the beta samples, calculated with one-second resolution. In our study, the recordings of one brain hemisphere determined the modulation of the stimulation amplitude delivered independently in both hemispheres (single-driver STN).

The programming of aDBS with the AlphaDBS device has been previously described^13,14^. In brief, the stimulation amplitude was adjusted for each patient within a predefined, clinically effective range for each STN (i.e., minimum amplitude (Amin) and maximum amplitude (Amax)), while the stimulation frequency and pulse width remained fixed. The two stimulation current limits were clinically defined as the Amin (providing 40-50% clinical benefit in meds-off state) and Amax (in the absence of side effects in the meds-on condition). aDBS stimulation proportionally changes within the range Amin-Amax according to the amplitude of the beta oscillation—when the beta oscillation increases, the stimulation amplitude increases; when the beta oscillation decreases, the stimulation amplitude decreases. The increase and decrease are bound within the two limits Amin and Amax through the definition of Pmin and Pmax (Figure 2A), representing the beta oscillation values at which the stimulation amplitude are set to Amin and Amax, respectively. These two beta values were defined during the calibration days of the study, based on the beta amplitude distribution. In terms of programming options, these beta values define the amount of time the patient is stimulated towards or at Amin or Amax.

**Figure 2.**
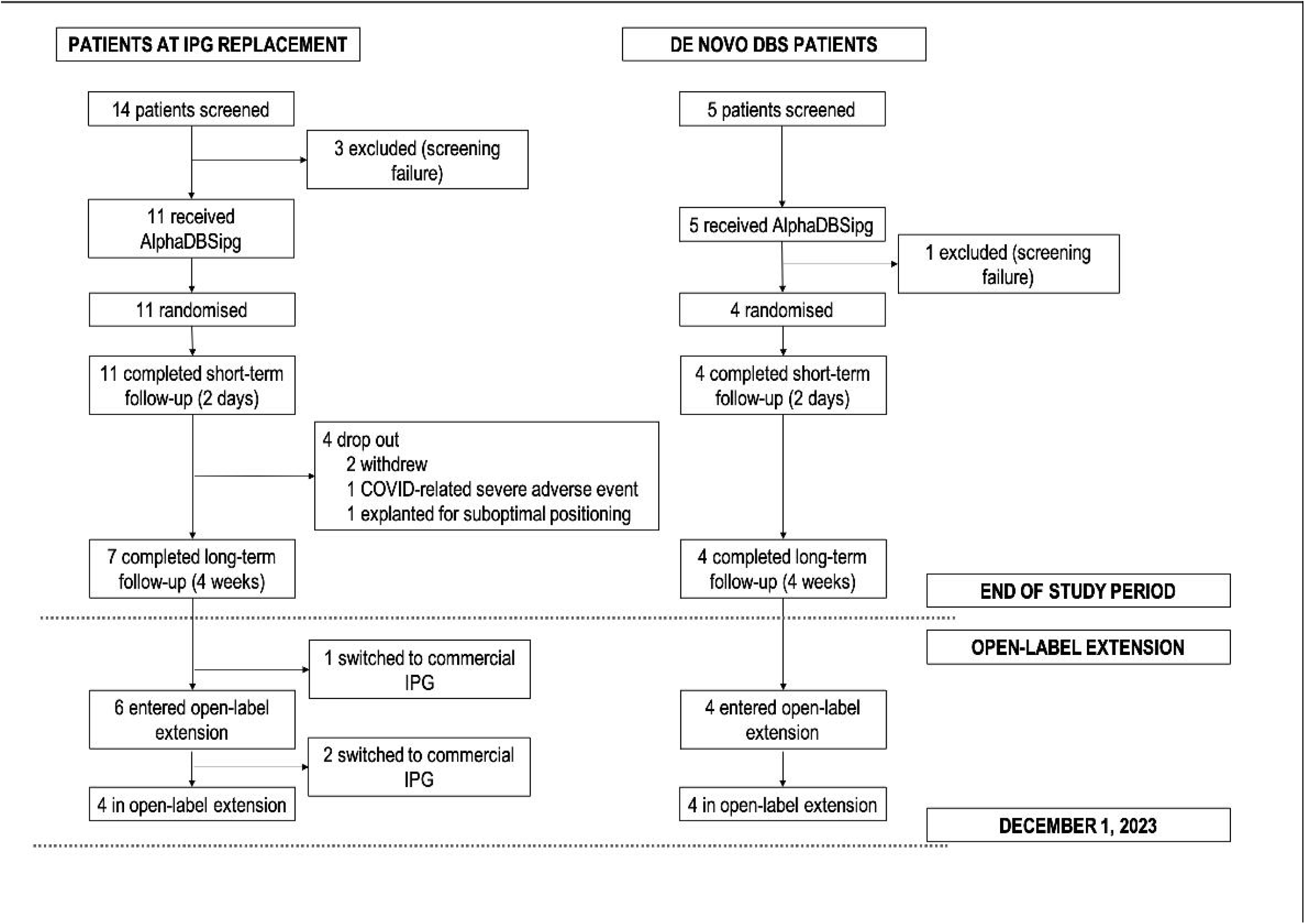
(A) Example of cDBS (left panel) and aDBS (right panel) programming from a representative patient. Each panel represent the histogram of the average distribution of beta power in the observation period (one week), during the daytime (yellow), nighttime (brown), and all-day (black line). The red and black lines represent DBS amplitudes in the left and right side respectively. Vertical dashed lines represent Pmin and Pmax, i.e., the values of beta power below/over which the DBS amplitude is set to Amin and Amax, respectively. Between Pmin and Pmax, DBS amplitude varies linearly according to the side specific setting. (B) Example of time-frequency spectra (5-35 Hz) collected by the AlphaDBS IPG during one representative week in cDBS (top panel) and aDBS (bottom panel). Abbreviations: Amin/max, minimum/maximum amplitude; (a/c)DBS, (adaptive/conventional)deep brain stimulation; IPG, implantable pulse generator; Pmin/max, minimum/maximum probability.

For safety reasons, we set Amax equal to cDBS in the first three patients (see Table 1). This programming limited the full exploitation of aDBS by preventing the delivery of stimulation amplitudes higher than cDBS for high beta power values. Programming in cDBS was performed as a standard of care ^20,21^. Figure 2B shows an example of LFPs recorded during one week of cDBS and aDBS.

**Table 1.** Stimulation settings.

| Patient N/Side/Active contact pair |  |  | Pulse (us) | Frequency (Hz) | cDBS Amplitude (mA) | aDBS Amin (mA) | aDBS Amax (mA) | aDBS Amax-cDBS Amplitude (mA) | Biomarker range (local field potentials) (Hz) |
| --- | --- | --- | --- | --- | --- | --- | --- | --- | --- |
| PATIENT 1 | Left | C+1- | 60 | 130 | 4 | 3.8 | 4 | 0 | 14-18 |
|  | Right | C+9- | 60 | 130 | 3.5 | 3.3 | 3.5 | 0 | 14-18 |
| PATIENT 2 | Left | C+1- | 60 | 130 | 2.4 | 2.2 | 2.4 | 0 | 12-20 |
|  | Right | C+9- | 60 | 130 | 1.8 | 1.6 | 1.8 | 0 | 12-20 |
| PATIENT 3 | Left | C+3- | 60 | 130 | 2.5 | 2 | 2.5 | 0 | 12-16 |
|  | Right | C+11- | 60 | 130 | 2.3 | 2 | 2.5 | 0 | 12-16 |
| PATIENT 4 | Left | C+1- | 60 | 130 | 2.3 | 2.1 | 2.5 | +0.2 | 12-20 |
|  | Right | C+9- | 60 | 130 | 2.8 | 2.6 | 3 | +0.2 | 12-20 |
| PATIENT 5 | Left | C+1- | 70 | 130 | 2.4 | 2.6 | 3.9 | +1.5 | 11-16 |
|  | Right | C+8- | 70 | 130 | 2.5 | 2.6 | 3 | +0.5 | 11-16 |
| PATIENT 6 | Left | C+1- | 60 | 130 | 2.5 | 2.4 | 3.4 | +0.9 | 11-16 |
|  | Right | C+9- | 60 | 130 | 2.5 | 2.4 | 3.2 | +0.7 | 11-16 |
| PATIENT 7 | Left | C+2- | 60 | 70 | 4 | 3.5 | 4.5 | +0.5 | 12-19 |
|  | Right | C+8- | 60 | 70 | 3.5 | 3 | 4 | +0.5 | 12-19 |
| PATIENT 8 | Left | C+2- | 60 | 130 | 2.2 | 1.8 | 2.8 | +0.6 | 12-18 |
|  | Right | C+9- | 60 | 130 | 2.1 | 1.8 | 2.8 | +0.7 | 12-18 |
| PATIENT 9 | Left | C+2- | 60 | 130 | 4 | 3.5 | 4.5 | +0.5 | N/A |
|  | Right | C+10- | 60 | 130 | 3.8 | 3.5 | 4 | +0.2 | N/A |
| PATIENT 10 | Left | C+2- | 60 | 130 | 3 | 2 | 3.3 | +0.3 | 12-20 |
|  | Right | C+10- | 60 | 130 | 3.1 | 1 | 3.1 | 0 | 12-20 |
| PATIENT 11 | Left | C+0- | 60 | 80 | 4 | 2.5 | 4.5 | +0.5 | 11-17 |
|  | Right | C+10- | 60 | 80 | 4 | 3 | 4.5 | +0.5 | 11-17 |
| PATIENT 12 | Left | C+2- | 60 | 130 | 2.4 | 2 | 2.9 | +0.5 | 11-20 |
|  | Right | C+9- | 60 | 130 | 2.3 | 2 | 3.5 | +1.2 | 11-20 |
| PATIENT 13 | Left | C+3- | 60 | 130 | 2 | 1.5 | 2.6 | +0.6 | 9-16 |
|  | Right | C+11- | 60 | 130 | 2.2 | 1.5 | 2.6 | +0.4 | 9-16 |
| PATIENT 14 | Left | C+3- | 60 | 130 | 2.3 | 1.5 | 3 | +0.7 | 12-20 |
|  | Right | C+10- | 60 | 130 | 1.8 | 1.2 | 1.8 | 0 | 12-20 |
| PATIENT 15 | Left | C+2- | 60 | 180 | 2.4 | 2 | 2.6 | +0.2 | 11-22 |
|  | Right | C+9- | 60 | 180 | 3.6 | 3 | 4 | +0.4 | 19-25 |

Abbreviations: Amin/max, minimum/maximum amplitude; (a/c)DBS, (adaptive/conventional)deep brain stimulation.

### Outcome measures

Our overall aim was to assess the safety, tolerability, and effectiveness of personalized, LFP-based aDBS that uses the linear proportional algorithm delivered by the AlphaDBS system. The primary objective was to evaluate the safety and tolerability of the AlphaDBS system when used in the cDBS and aDBS mode. The safety and tolerability was based on the occurrence of device-related adverse events (AEs). The efficacy of aDBS was evaluated through the following secondary measures:

- PD-related motor symptoms (i.e., bradykinesia, rigidity, and tremor at rest) and their fluctuations through repeated clinical assessments (UPDRS-III).
- Dyskinesia and their fluctuations through repeated clinical assessments (UDysRS, patient diary).
- GOT, defined as the time spent during the day in ON without troublesome dyskinesia, measured using a three-day patient diary ^19^.
- Patient preference, measured as the stimulation mode preferred by the patient while still blinded at the end of the LT-FUP.

### Statistical analysis

Descriptive statistics were computed for all clinical endpoints (UPDRS-III, UDysRS, GOT) in each stimulation mode. Throughout the text, data are presented as mean±standard deviation (SD). Correlations between outcome variables and demographic (sex, age) and clinical (disease duration, levodopa equivalent daily dose pre-surgery) variables were performed.

Even though this first-in-human study was not designed to claim effectiveness of aDBS or superiority of aDBS over cDBS, an exploratory analysis was conducted to compare the two stimulation modes and better understand their effects on patients in chronic conditions. To avoid limitations due to the small sample size and to the possibility of sequential testing, in addition to classical frequentist statistics we decided to use the Bayesian statistical approach. This estimates the Bayes factor (BF10), representing the ratio between the likelihood of the alternative hypothesis and that of the null hypothesis to explain the observed data, thus giving a level of evidence in favor of the alternative hypothesis (BF10>1) or of the null hypothesis (BF10<1). According to standard practices, the levels of evidence adopted were anecdotal (1/3<BF10<3), moderate (<1/3 or >3), strong (<1/10 or >10), very strong (<1/30 or >30), or decisive (<1/100 or >100). The JASP software (version 0.16.3, University of Amsterdam, NL) was used for the analysis ^22^.

As stated in the protocol description ^17^, a sample size of 15 patients was considered adequate to confirm safety and tolerability of aDBS. In addition, a population of 15 patients would allow the non-inferiority of the mean change in UPDRS-III score (ΔUPDRS-III = UPDRS-III score collected at screening after pausing all medications versus UPDRS-III score collected after two weeks of stimulation) to be tested in aDBS versus cDBS. We considered a non-inferiority margin equal to the minimal clinically important difference (MCID) proposed in the literature ^23^, assuming a standard deviation of 4.3 points ^24^ and a type I error probability equal to 0.05 and power of 80% in the frequentist approach. We also estimated the non-inferiority using the Bayesian one-sample t-test.

In addition to the previously defined population analyses, we also explored the intra-patient response to aDBS by assessing patient responses reaching the MCID ^25^. This approach is grounded in the definition of a threshold on clinical scales representing a difference that is perceived as relevant by patients. In PD, the literature provides the MCID estimation for all of the clinical endpoints used in our study (UPDRS-III: 4.83 points ^23^; UDysRS: 3.9 points ^26^; GOT: 2 hours ^27^). To assess whether the difference between the response of a single patient to cDBS and aDBS on a certain scale was clinically meaningful, we calculated the difference between the value of the endpoint obtained by a patient with aDBS and cDBS, and compared it to the MCID. A value above the MCID represented a clinically relevant difference in favor of one of the two modes, whereas a difference below the MCID was considered to be no-difference (equality of response). We then calculated the percentage of patients that reached the MCID in favor of aDBS or cDBS.

## RESULTS

### Patients

Fifteen patients were enrolled (19 screened, Figure 3). The details of enrolled patients, their baseline characteristics, and their follow-ups are provided in Table 2.

**Figure 3.**
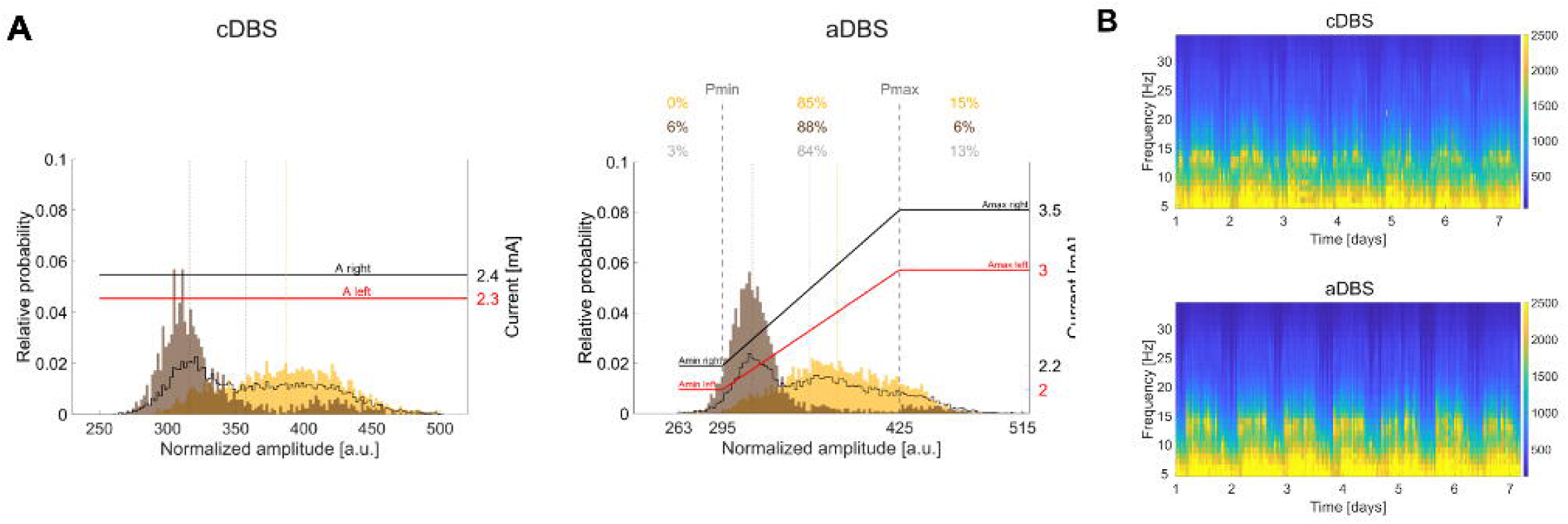
Study profile. The study profiles of the IPG replacement population (left side) and of the de novo DBS population (right side) are represented. Abbreviations: DBS, deep brain stimulation; IPG, implantable pulse generator.

**Table 2.**
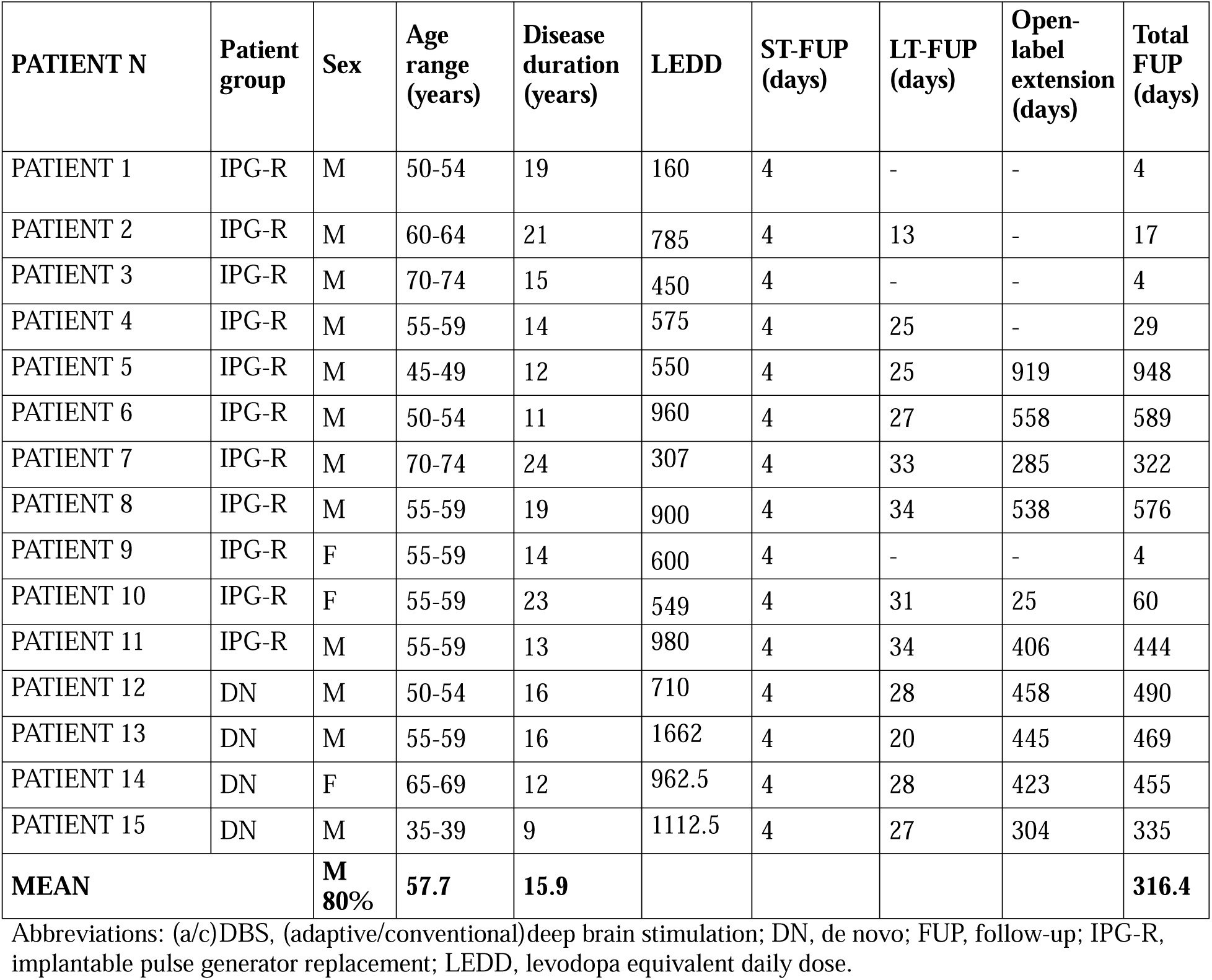
Demographic and clinical details at screening visit.

In the IPG replacement group, 14 patients were screened. Of them, three were excluded (screen failure) because the LFPs were confounded by artifact (mostly cardiac artifact). The remaining 11 patients were enrolled, randomized, and completed the ST-FUP. Seven also completed the LT-FUP. Of the four patients that did not complete the LT-FUP, two dropped out for personal reasons not related to the study, one was explanted after ST-FUP for suboptimal positioning of the AlphaDBS IPG that prevented it from correctly recharging, and one dropped out during the LT-FUP due to a serious AE not related to the study procedure or device (i.e., pulmonary embolism due to COVID-19 pandemic). Of the seven patients who completed the study, six decided to keep the investigational device at the end of the trial (open-label extension phase). One patient asked for a non-rechargeable device.

In the de novo group, five patients were screened. Of them, one was excluded (screening failure) because the LFPs were confounded by cardiac artifact. This patient did not enter the study (not randomized) but kept the AlphaDBS IPG in the cDBS mode, and was included in the safety analysis. The remaining four patients completed both the ST-FUP and LT-FUP. All patients in the de novo group decided to keep the investigational device at the end of the trial (open-label extension phase). Overall, the total available follow-up (to December 2023) was 4,746 days, which corresponds to about 158 months. The mean follow-up was 316 days (corresponding to 10 months).

### aDBS safety

No AEs related to the administration of aDBS were recorded during the study period. Two serious AEs unrelated to the DBS mode were recorded, one being a pulmonary embolism related to COVID-19 and the other related to a technical failure of device recharging. Other mild, device-related AEs were reported, which were independent of the stimulation type.

The daily average of the total electrical energy delivered (TEED) was similar when the patients were treated in aDBS and cDBS mode during the LT-FUP (aDBS versus cDBS: 114.62 ± 67.72 versus 106.11 ± 42.31 μW). The large variability was dependent on differences in individualized programming (Table 1).

### aDBS versus cDBS on motor symptoms

Correlations between the outcomes measured and the main demographic and clinical variables are reported in Table 3. In summary, both aDBS and cDBS were similarly effective for motor symptoms during the ST-FUP, with aDBS improving dyskinesia the most. According to the intra-patient analysis, aDBS showed better outcomes in most patients during the LT-FUP, with 82% (9 out of 11) choosing (blinded) aDBS at the end of the study. For most of the variables during the ST-FUP and LT-FUP, we found non-significant correlations. Of note, Bayesian statistics for correlations provided only an anecdotal level of evidence against the alternative hypothesis (1/3<BF<1). The only significant correlation was found in the aDBS GOT with respect to sex, even though the sample size was too unbalanced to draw a conclusion (female n=2, male n=9).

**Table 3.** Correlations between clinical outcomes and clinical characteristics.

|  |  | Sex, mean (SD) |  |  | Avg (SD) | Age | Disease Duration |
| --- | --- | --- | --- | --- | --- | --- | --- |
|  |  | Female | Male | Wilcoxon p Bayesian BF<sub>10</sub> |  | Spearman's p Bayesian BF<sub>10</sub> | Spearman's p Bayesian BF<sub>10</sub> |
| **ST-FUP**<br>(N=15;<br>M:F 12:3) | **Delta UPDRS III MedOFF/StimOFF F - MedOFF/StimON Alpha cDBS** | 26.0<br>(16.1) | 34.3<br>(17.2) | 0.47<br>0.59 | 32.6<br>(16.7) | 0.44<br>0.34 | 0.48<br>0.49 |
|  | **Delta UPDRS III MedOFF/StimOFF F - MedOFF/StimON Alpha aDBS** | 20.0<br>(18.0) | 34.3<br>(17.3) | 0.22<br>0.80 | 31.5<br>(17.7) | 0.35<br>0.48 | 0.97<br>0.36 |
|  | **Delta UPDRS III MedOFF/StimOFF F - MedON/StimON Alpha cDBS** | 35.3<br>(13.5) | 41.2<br>(21.1) | 0.82<br>0.53 | 40<br>(19.5) | 0.29<br>0.71 | 0.51<br>0.36 |
|  | **Delta UPDRS III MedOFF/StimOFF F - MedON/StimON Alpha aDBS** | 33.3<br>(13.7) | 35.9<br>(19.2) | 1<br>0.51 | 35.4<br>(17.8) | 0.13<br>0.32 | 0.32<br>0.68 |
|  | **Max UDysRS cDBS** | 12<br>(10.4) | 7.3<br>(7.4) | 0.46<br>0.64 | 8.2<br>(7.9) | 0.08<br>0.90 | 0.18<br>0.48 |
|  | **Max UDysRS aDBS** | 8.7<br>(7.8) | 4.5<br>(5.4) | 0.41<br>0.71 | 5.3<br>(5.9) | 0.92<br>0.33 | 0.90<br>0.33 |
| **LT-FUP**<br>(N=11;<br>M:F 9:2) | **UPDRS III MedON/StimON cDBS** | 20.5<br>(7.8) | 23<br>(7.9) | 0.69<br>0.58 | 22.5<br>(7.5) | 0.72<br>0.39 | 0.49<br>0.45 |
|  | **UPDRS III MedON/StimON aDBS** | 24.5<br>(2.1) | 17<br>(7.4) | 0.12<br>0.89 | 18.4<br>(7.3) | 0.60<br>0.40 | 0.99<br>0.38 |
|  | **GOT cDBS** | 8.3<br>(0.4) | 12.2<br>(4.4) | 0.15<br>0.81 | 11.5<br>(4.2) | 0.79<br>0.38 | 1<br>0.37 |
|  | **GOT aDBS** | 4.9<br>(1.3) | 15.0<br>(2.8) | 0.05<br>23.0 | 13.2<br>(4.8) | 0.30<br>0.86 | 0.91<br>0.42 |

Abbreviations: (a/c)DBS, (adaptive/conventional)deep brain stimulation; GOT, good on time; LT/ST-FUP, long-term/short-term follow-up; SD, standard deviation; UDysRS, Unified Dyskinesia Rating Scale; UPDRS, Unified Parkinson's Disease Rating Scale.

The ST-FUP results (n=15, Table 2) confirmed an equivalent improvement in the UPDRS-III between cDBS and aDBS—supporting the concept that aDBS (alone or in combination with levodopa) significantly improves PD motor symptoms at least as well as cDBS. When the patient received stimulation only (meds-off/stim-on condition), the average improvement in UPDRS-III was 56% (32.6 points, (standard deviation (SD) 16.7) and 54% (31.5 points, SD 17.7) with cDBS and aDBS, respectively (Table 3). When the patient received both stimulation and medical therapy (meds-on/stim-on condition), the improvement in the UPDRS-III increased to 66% (40 points, SD 19.5) and 61% (35.4 points, SD 17.9) with cDBS and aDBS, respectively.

Considering individual responses, we observed that eight patients had an equal improvement with aDBS and cDBS (difference aDBS versus cDBS <1 MCID), two patients had greater improvement with aDBS, and five patients had greater improvement with cDBS in the meds-off/stim-on condition. In the meds-on/stim-on condition, four patients had an equal improvement with aDBS and cDBS (difference aDBS versus cDBS <1 MCID), four patients had greater improvement with aDBS, and seven patients had greater improvement with cDBS. When considering only patients in the IPG replacement group, the same trend was confirmed when the ability of aDBS to control motor symptoms was compared with that of the pre-surgical cDBS (aDBS versus pre-surgery cDBS, meds-off/stim-on: 56% ± 15% versus 53% ± 9%).

In general, patients showed very mild dyskinesias during the assessments conducted during the ST-FUP. However, comparing the worst UDysRS score obtained across all assessments conducted in aDBS and cDBS mode, we observed some additional reduction in dyskinesia in the aDBS mode (cDBS vs aDBS: 8.2±7.9 vs 5.3±5.9). We also considered individual responses for the UDysRS score and observed that eight patients had no clinically significant difference between the worst UDysRS recorded in aDBS and cDBS modes; a clinically significant lower UDysRS was noted in six patients with aDBS (UDysRS aDBS – cDBS: −3.9 points) and one with cDBS (UDysRS aDBS – cDBS: 3.9 points).

During the LT-FUP (n=11), aDBS tended to provide better improvement than cDBS (Figure 4A and B). More specifically, when considering the UPDRS-III (meds-on/stim-on) after two weeks of at-home treatment, aDBS showed better average scores than cDBS (aDBS versus cDBS: 18.36 ± 7.32 versus 22.50 ± 7.5, Figure 4B). This corresponded to an average change of 53% for cDBS versus 61% for aDBS, even though the superiority of aDBS did not reach statistical significance (p=0.06). When analyzing the non-inferiority of aDBS versus cDBS (ΔUPDRSIII aDBS - ΔUPDRSIII cDBS), using the MCID of 4.83 as non-inferiority margin, aDBS was significantly non-inferior to cDBS (Wilcoxon p=0.005, Bayesian BF_0-_ factor =26.35, strong evidence).

**Figure 4.**
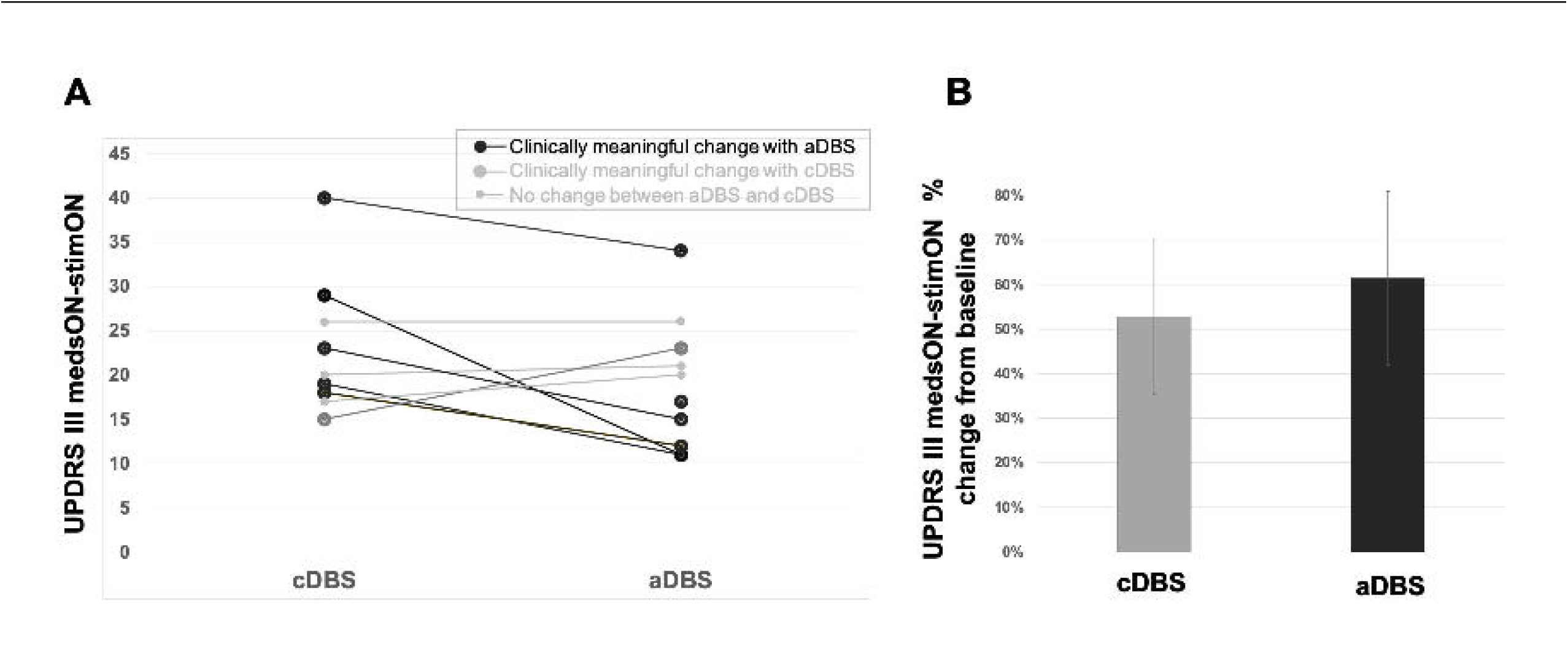
A: Individual values of the UPDRS III recorded after two weeks of cDBS and aDBS. Big dots represent clinically meaningful changes in favor of aDBS (black) or cDBS (grey). B: Average improvement in UPDRS III scores in medsOn-stimOn condition after two weeks of stimulation compared to baseline. Error bars are standard deviations. Abbreviations: (a/c)DBS, (adaptive/conventional)deep brain stimulation; UPDRS, Unified Parkinson’s Disease Rating Scale.

Evaluation of individual responses during the LT-FUP (Figure 4A) demonstrated that six patients experienced a clinically significant higher improvement in motor symptoms with aDBS (UPDRS-III aDBS versus cDBS >1 MCID), with a difference in UPDRS-III ranging from 1 MCID (4.83 points) to 3 MCID (14.49 points). Of the other four patients, three had an equal improvement with aDBS and cDBS and one had a clinically significant higher improvement in motor symptoms with cDBS.

In the LT-FUP, patients were asked to complete a three-day diary. Patients experienced a very high percentage of GOT (on time with non-troublesome dyskinesias) after the two follow-up weeks, both in cDBS (11.46 ± 4.24 hours, corresponding to 72% of waking time) and aDBS (13.15 ± 4.8 hours, corresponding to 81% of awake time) (Figure 5A-C). Considering the intra-patient change in GOT (ΔGOT = GOTaDBS - GOTcDBS) and categorizing such differences into groups that corresponded to multiples of the MCID (two hours), we found that three patients experienced a clinically significant difference in GOT with aDBS, ranging from 1 MCID (+ two hours) to >3 MCID (> six hours), five had no clinically significant difference between aDBS and cDBS (difference within the two hours), while three had a clinically significant difference in GOT with cDBS (Figure 5D). Interestingly, the patients with a clinically significant difference in GOT with aDBS were not the same as those who had a clinically significant difference in chronic UPDRS-III with aDBS.

**Figure 5.**
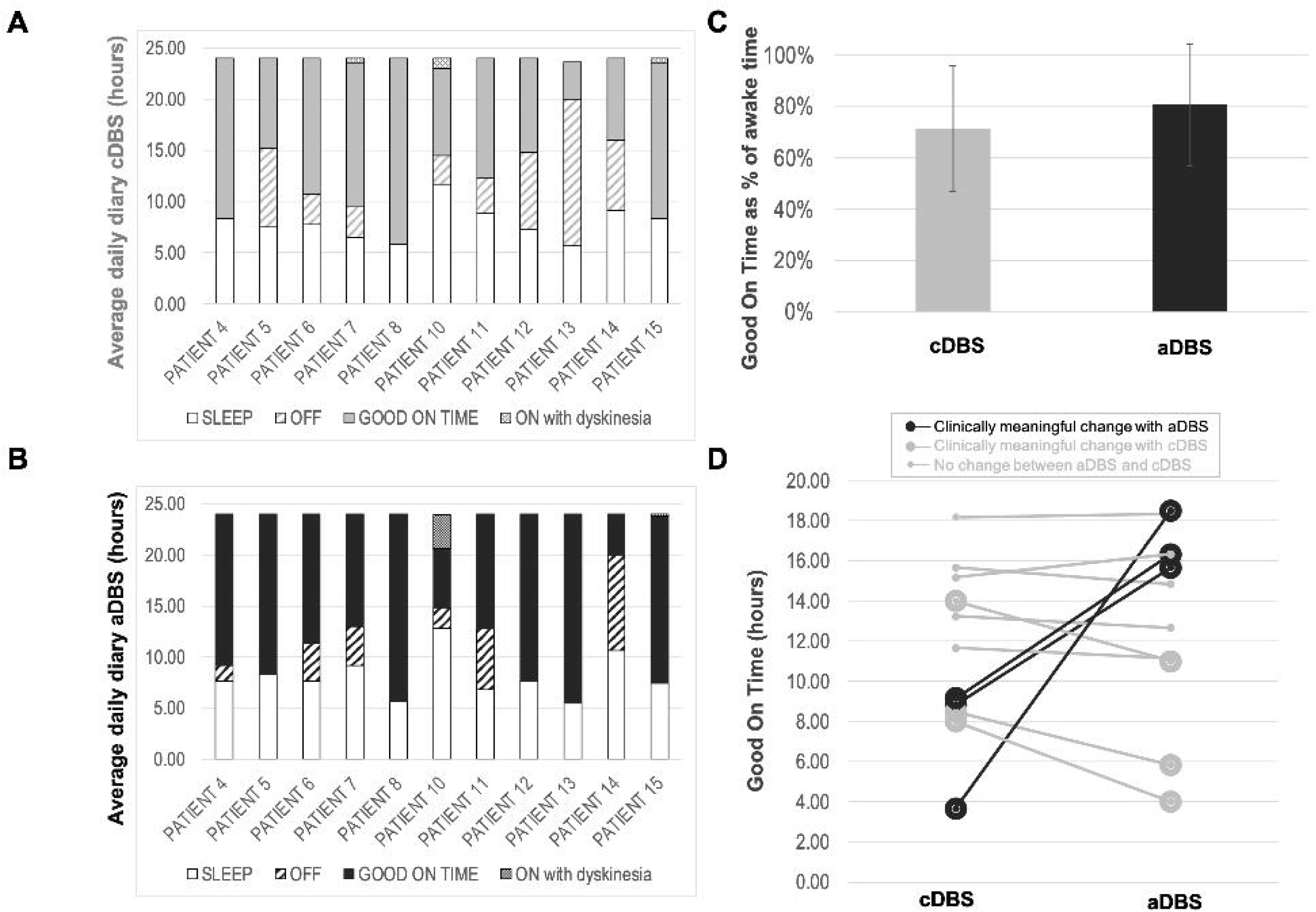
Average of three-day patient diary data collected after two weeks in A: cDBS mode and B: aDBS mode. The waking time was classified as OFF time, Good On Time, and On Time with troublesome dyskinesia. C: Comparison of GOT in cDBS and aDBS, expressed as proportion (%) of waking time. Error bars are standard deviations. D: Individual values of the average GOT (hours) in cDBS and aDBS mode. Big dots represent clinically meaningful changes in favor of aDBS (black) or cDBS (grey). Abbreviations: (a/c)DBS, (adaptive/conventional)deep brain stimulation; GOT, good on time.

### Patient preference

Of the ten patients who decided to keep the AlphaDBS system at the end of the study, eight (80%) preferred the aDBS mode. As shown in Table 4, patients preferring aDBS experienced a significant improvement measured either by the UPDRS-III or by GOT. When pooled together, individual responses accounted for 80% of patients significantly responding more to aDBS than cDBS. In addition, anecdotal reports from patients themselves or from their caregivers highlighted improvements in clinical domains that were not fully assessed in the patient diaries or clinical scales (e.g., speech and gait) ^14^.

**Table 4.** Patient preference.

| Patient | UPDRS III medsON/stimON |  |  | Good On Time (GOT) |  |  | Patient's preference |
| --- | --- | --- | --- | --- | --- | --- | --- |
|  | cDBS %improvement from baseline | aDBS % improvement from baseline | Clinically meaningful improvement aDBS vs cDBS | GOT cDBS (hours) | GOT aDBS (hours) | Clinically meaningful improvement aDBS vs cDBS |  |
| PATIENT 4 | 25% | 36% | aDBS = cDBS | 15.67 | 14.83 | aDBS = cDBS | **aDBS** |
| PATIENT 5 |  | 78% |  | 8.83 | 15.67 | aDBS | **aDBS** |
| PATIENT 6 | 70% | 83% | aDBS | 13.25 | 12.67 | aDBS = cDBS | **aDBS** |
| PATIENT 7 | 68% | 79% | aDBS | 14.00 | 11.00 | cDBS | **aDBS** |
| PATIENT 8 | 53% | 82% | aDBS | 18.17 | 18.33 | aDBS = cDBS | **aDBS** |
| PATIENT 10 | 52% | 52% | aDBS = cDBS | 8.50 | 5.83 | cDBS | **aDBS** |
| PATIENT 11 | 63% | 75% | aDBS | 11.67 | 11.17 | aDBS = cDBS | **aDBS** |
| PATIENT 12 | 53% | 51% | aDBS = cDBS | 9.17 | 16.33 | aDBS | **aDBS** |
| PATIENT 13 | 21% | 48% | aDBS | 3.67 | 18.50 | aDBS | **aDBS** |
| PATIENT 14 | 53% | 28% | cDBS | 8.00 | 4.00 | cDBS | **cDBS** |
| PATIENT 15 | 70% | 64% | cDBS | 15.17 | 16.33 | aDBS = cDBS | **cDBS** |

Abbreviations: (a/c)DBS, (adaptive/conventional)deep brain stimulation; GOT, good on time; UPDRS, Unified Parkinson's Disease Rating Scale.

Patients who entered the open-label extension phase experienced persistent improvement after implantation, with an average of 96% GOT of awake time reported in the last available clinical diaries, completed as part of the routine clinical patient visits (Figure 6). For three patients, data were collected more than 12 months after surgery; in the remaining four patients, data were collected after more than six months after surgery. One patient who had entered the open-label extension for less than one month was not included in the analysis.

**Figure 6.**
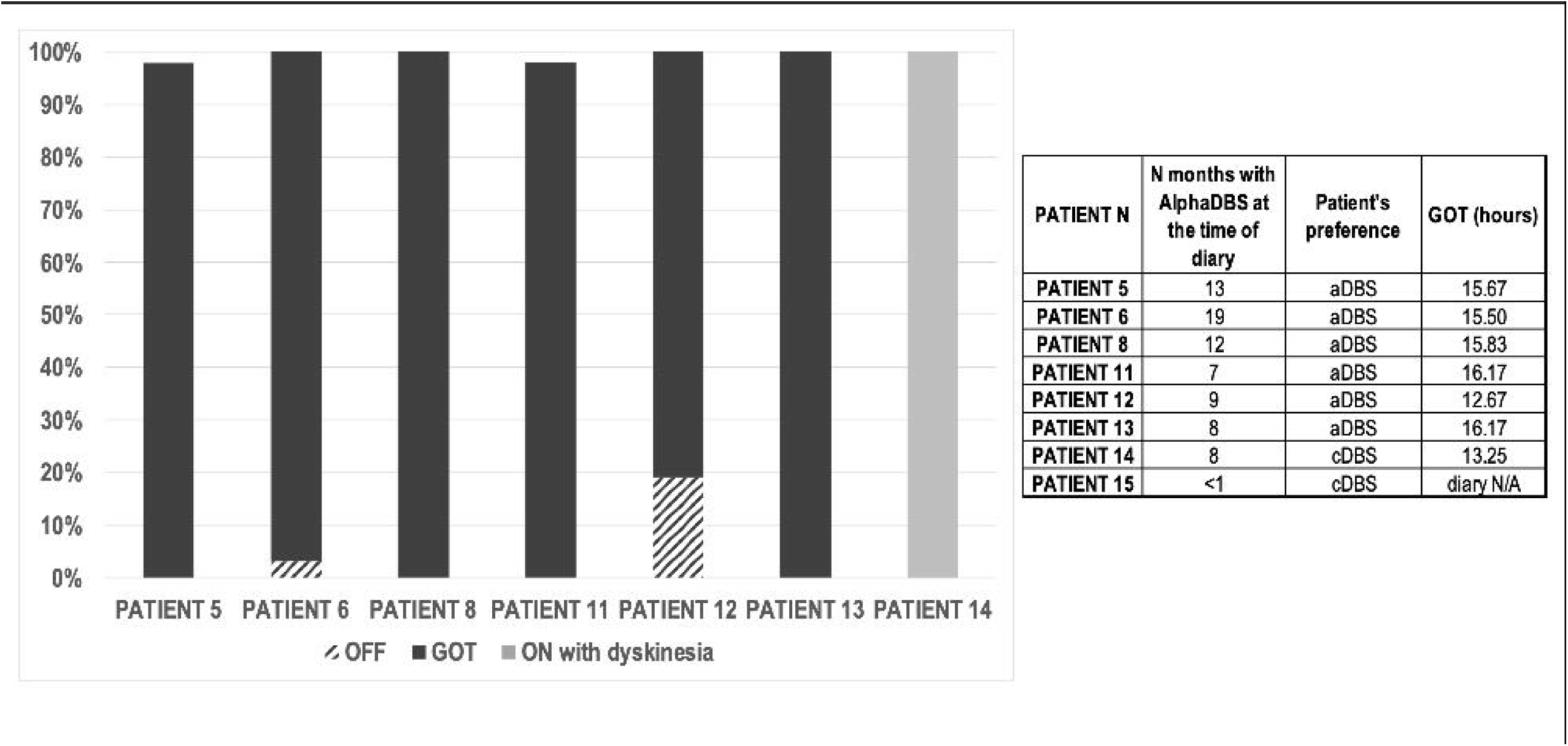
Analysis of patient diary data collected during the last available routine visit. The waking time was classified as OFF time, Good On Time, and On Time with troublesome dyskinesia (corresponding to severe dyskinesia). Patient stimulation mode and months of follow-up are reported in the table. Abbreviations: (a/c)DBS, (adaptive/conventional)deep brain stimulation; GOT, good on time.

## DISCUSSION

Our results indicate that aDBS is safe and in some patients may provide additional clinical improvement over cDBS. Most patients chose to continue treatment in the adaptive mode. In the longer term (after six months in the open-label extension), patients with aDBS showed significant and stable improvement (>90% good on time as a percentage of awake time). Ours is one of the largest studies to date to compare aDBS with cDBS ^8,10,28,29^; a larger study has been recently concluded (ADAPT-PD trial, with more 60 patients) ^11^.

It is important to note that our study consecutively enrolled patients eligible for STN-DBS without further specific selection criteria (e.g., severity of motor fluctuations, dosage or number of levodopa intakes, etc.) that would have eventually maximized the benefits from aDBS. This choice allowed us to study aDBS in a real-world environment, highlighting its benefits but also limitations.

In our study, aDBS and cDBS were randomly applied, thus minimizing the confounding effect of sequential application. This approach differs from the ADAPT-PD trial, in which cDBS was always administered first ^11^. The choice between randomization and sequential administration has a relevant impact on the study: while the sequential approach uses the cDBS response to inform decisions about aDBS, thus optimizing aDBS programming, the randomization approach avoids possible confounding factors due to the previous therapy.

In our study, aDBS performed at least as well as cDBS in controlling motor symptoms; in the majority of patients, aDBS achieved better results in terms of clinically relevant improvement. This outcome was recorded during the LT-FUP and the long-term, open-label extension (using data from clinical diaries). This is important information for future studies with aDBS. Home monitoring may be particularly important in describing the real benefit of stimulation programs that modulate stimulation delivery over long periods, and thus may be less frequently captured by punctual in-person neurological assessment.

The improvement in GOT observed during the LT-FUP was similar to that recently reported in another study ^28^. Although this study was conducted in only four patients and used a different aDBS paradigm, the authors reported a decreased time spent with most the bothersome symptom with aDBS versus cDBS.

It is important to note that cDBS provided very good symptoms relief, thus confirming that cDBS programming has also been optimized. The improvement margins with aDBS were therefore limited. Thus in our analysis, we decided to use the concept of the MCID to assess the intra-individual response to aDBS and cDBS. The MCID represents a clinically meaningful difference that not only includes the magnitude of the improvement, but also the value of the change as perceived by the patient ^25^.

A relevant finding is the preference of our patients for aDBS over cDBS (80%) that remained stable over time up to two years. In this time frame, aDBS continued performing very well and, despite the ability to switch to cDBS at any time, remained the chosen treatment. Interestingly, the percentage of patients who chose aDBS over cDBS (80%) was higher than the percentage of patients showing a clinically relevant difference between aDBS and cDBS for the UPDRS-III score (6 out of 11) and the GOT (3 out of 11) at LT-FUP. However, when pooling the results of these two measures of clinical performance, we observed that patients preferring aDBS had at least one evaluation in which the difference between aDBS and cDBS was clinically relevant (>1 MCID).

The fact that not all patients responded similarly at different scales is an important observation. As previously underlined, our study design was not selected to emphasize aDBS performance and therefore the benefits of aDBS were captured in different clinical domains. This observation suggests that the concept of “response” to aDBS is multifactorial and deserves further research to be fully exploited.

Even though the ST-FUP may be of limited interest for the chronic application of aDBS, we decided to include this first assessment in our study with the aim of verifying the patient response to both the AlphaDBS device when implanted and aDBS in a controlled environment. Indeed, the results of the ST-FUP revealed that with aDBS, levodopa-related dyskinesias tended to be more controlled. This finding is in line with previous studies using an externalized IPG, conducted in the acute postoperative phase ^12,24,30,31^.

Safety is another aspect that in our study was in line with previous studies. Although AEs were reported, none were related to the aDBS mode per se, but mainly related to technical issues (mostly concerning battery recharging).

In contrast to previous literature, the TEED in aDBS in our study was equal to or higher than in cDBS. Previous studies advocated a lower TEED in aDBS to possibly reduce the risk of AEs from chronic cDBS ^12,32,33^. However, we should consider that a higher TEED is delivered transiently and only when clinically necessary in aDBS. This observation opens a new perspective on aDBS, not only as an “energy saving” strategy, but also as an enhancement of the positive effect of stimulation.

The neurologists involved in our study were free to decide how to program the AlphaDBS device in each mode. It should be mentioned that the first three patients were programmed conservatively, limiting the Amax to the current delivered in the cDBS. This precautionary measure was not applied to subsequent patients, to allow us to better exploit aDBS. Note that in the open-label extension phase, it was often necessary to redefine the beta minimum and maximum thresholds—an easy task because of the minute-by-minute storage of patient-specific beta power (Figure 2), which allows direct correspondence of the biomarker with the clinical state of the patient.

In conclusion, our pilot study in PD patients showed that aDBS is safe and effective, even after years of administration, and should be an available modality for all PD patients receiving DBS. In addition to the possibility of greater clinical benefit and hopefully fewer AEs from chronic cDBS, the large amount of data recorded with these devices opens up new possibilities to better understand the pathophysiology of neurological disorders and the mechanisms of action of DBS.

## Data Availability

All data produced in the present study are available upon reasonable request to the authors

